# A systematic review of efficacy and safety of intramuscular midazolam versus midazolam by other routes or other non-intravenous benzodiazepines in children with convulsive status epilepticus

**DOI:** 10.64898/2025.12.03.25341538

**Authors:** Richard F. M. Chin, Michael J. Vasey, Frank M. C. Besag

## Abstract

**Background:** Convulsive status epilepticus (CSE) is the most common neurological emergency in children. Without prompt, safe and effective intervention, CSE can cause adverse neurological sequelae or death. There is increasing use of intramuscular midazolam for the treatment of childhood CSE.

The aim of this systematic review is to investigate the efficacy and safety of intramuscular midazolam versus midazolam by other routes or other non-intravenous benzodiazepines in childhood CSE.

**Methods:** MEDLINE, PubMed, Embase, Cochrane CENTRAL and Google Scholar were searched from 01/01/1980 to 29/08/2025. The primary outcome of interest was cessation of seizures within 5-10 minutes of treatment. Secondary outcomes included time to seizure cessation and incidence of respiratory depression/hypotension.

**Results:** Five studies were identified comparing intramuscular midazolam with buccal midazolam, intranasal midazolam or rectal diazepam. In single studies, there was no difference between intramuscular midazolam and intranasal midazolam or rectal diazepam in achieving seizure cessation, but intramuscular midazolam was associated with shorter time to seizure cessation. Meta-analysis was only possible comparing intramuscular midazolam with buccal midazolam. Intramuscular midazolam was superior to buccal midazolam for time to seizure cessation (p<0.00001). While there were no inter-group differences in other outcomes, point estimates favoured intramuscular midazolam. Few cases of respiratory depression or hypotension were reported with no inter-group differences. No studies involved treatment by paramedics, sample sizes were modest, and none were in Western high-income countries. Quality of evidence was low or very low.

**Discussion:** There is insufficient clinical trial evidence for safety and efficacy of intramuscular midazolam compared to midazolam by other routes or other non-intravenous benzodiazepines for childhood CSE.

**Key points:**

- Out-of-hospital treatment is usually needed to achieve prompt seizure cessation in convulsive status epilepticus.
- Limited data indicate that intramuscular midazolam achieves faster seizure cessation than buccal midazolam.

## 1. Introduction

Convulsive status epilepticus (CSE) is operationally defined for treatment purposes as a tonic, clonic, or tonic-clonic seizure of at least five minutes duration or two or more such seizures in succession without recovery of consciousness between seizures [1, 2]. Seizures that have not spontaneously resolved or been successfully aborted within five minutes are more likely to continue for at least 30 minutes, after which there is a substantial risk of serious long-term neurological and neurodevelopmental sequelae, including neuronal injury, neuronal death, alteration of neuronal networks and functional deficits [2], and death. However, the time to initiation of emergency antiseizure medication (ASM) after onset of CSE is often greater than 30 minutes, especially where treatment is not commenced until admission to the hospital emergency department [3, 4]. Prognosis worsens with longer duration of continuous seizure activity, particularly where this exceeds 1-2 hours [5].

CSE is one of the most common medical neurological emergencies in children, estimated at between 17 and 23 episodes per 100 000 individuals per year in developed countries, and often occurs in children with no prior history of seizures or epilepsy [1, 6, 7]. Incidence is higher than in adults [8], highest in children under the age of five years [9], and decreases with increasing age [1]. Although mortality rates are lower than in adults [10], long-term impairments and increased mortality after childhood CSE are common [11]. The cumulative incidence of subsequent epilepsy at nine-years post-CSE is around 25% and neurological, cognitive and behavioural impairments are reported in at least one third of individuals. Long-term mortality rates are between 5% and 17% [12].

Since seizures in this population are more likely to occur either at home or in the community, and transfer to hospital may introduce avoidable delays in the initiation of treatment, there is considerable interest in optimising out-of-hospital interventions that can be administered quickly and effectively by both non-medical personnel and paramedics.

Benzodiazepines are the standard first-line emergency treatment for CSE [9, 13]. However, ongoing seizures become progressively less responsive to these drugs, possibly due to cellular internalisation (“receptor trafficking”) of the target inhibitory gamma-aminobutyric acid (GABA)_A_ receptors as an effect of continuous seizure activity [14, 15]. Rapid, effective and safe intervention for CSE is therefore essential to mitigate the risk of adverse short-term and longer-term outcomes [14].

The current guidelines of the UK National Institute for Health and Care Excellence (NICE) recommend either buccal midazolam (unlicensed for adults) or rectal diazepam for community intervention where intravenous access is unlikely to be possible, or intravenous lorazepam where intravenous access and resuscitation facilities are immediately available [16].

Midazolam is a rapidly absorbed, short-acting, water-soluble benzodiazepine [17, 18] which, as an alternative to the buccal route, can be administered intranasally, intramuscularly or intravenously. Buccal midazolam and intranasal midazolam have shown efficacy for cessation of childhood CSE at least comparable to that of rectal diazepam, are considered easier to administer than rectal diazepam and are more socially acceptable [19–27]. There are fewer data for intramuscular midazolam in out-of-hospital paediatric use; however, a report by the Guideline Committee of the American Epilepsy Society concluded that there was Level A evidence to support the use of intramuscular midazolam in adult CSE and Level B evidence for its use in paediatric CSE [28]. The most recent Joint Royal Colleges Ambulance Liaison Committee (JRCALC) guidelines specify that intramuscular midazolam or buccal midazolam are preferred first-line paramedic treatment given their superiority to rectal diazepam.

The objective of this systematic review is to examine the existing data for the efficacy and safety of intramuscular midazolam compared with midazolam administered by any other route and other non-intravenous benzodiazepines in the treatment of CSE in children either out of hospital or in-hospital, including in the accident and emergency department.

## 2. Methods

### 2.1 Search and eligibility criteria

The review was conducted in accordance with the PRISMA (Preferred Reporting Items for Systematic Reviews and Meta-Analyses) guidelines (prisma-statement.org). The review protocol was registered with PROSPERO (CRD420251129713).

A systematic literature search was conducted in MEDLINE, PubMed, Embase, Cochrane CENTRAL and Google Scholar for human, non-placebo controlled, comparative clinical trials published between 01/01/1980 and 29/08/2025. The detailed search strategy is available in the supplementary material. The reference lists of relevant reviews and articles were manually screened to identify additional studies. Randomised and quasi-randomised controlled trials, irrespective of blinding, were included. Studies were eligible if they reported data for children aged 0-18 years who had an acute convulsive seizure, treated with one or more ASM either in or out of hospital, irrespective of the cause or duration of the seizure, or whether the child had a known existing seizure disorder or the seizure was the first in their lifetime; and compared seizure outcomes between children treated with intramuscular midazolam and children treated with midazolam administered by another, non-intravenous, route or another non-intravenous benzodiazepine. There were no language or other restrictions.

The primary outcome of interest was cessation of the seizure within 10 minutes of commencement of drug administration. Secondary outcomes included time to clinical seizure cessation from the time of drug administration; recurrence of seizures within one hour of initial seizure cessation; and incidence of respiratory depression or hypotension.

Search results were imported to Covidence (covidence.org) and duplicate articles identified and excluded. The titles and abstracts of the remaining articles were independently screened by two of the authors (RC and MV). The full texts of potentially relevant studies were assessed against the eligibility criteria and disagreements resolved by consensus.

Data from studies meeting the eligibility criteria, including study design, participants, methodology, inclusion and exclusion criteria, outcomes, results and source of funding or other support, were independently extracted using a bespoke data form created in Covidence and discrepancies identified and resolved by consensus. Missing data were sought from the authors of the original studies. Data were exported to RevMan Web (revman.cochrane.org) for analysis.

Risk of bias in the included studies was assessed using the Cochrane Collaboration Risk of Bias Tool Version 2 (RoB2) and overall study quality assessed using the Grading of Recommendations, Assessment, Development and Evaluation (GRADE) approach (gradepro.org).

### 2.2 Data synthesis

Primary data analysis was performed on the basis of intention-to-treat. Heterogeneity between studies was assessed using the restricted maximum-likelihood (REML) method and visual inspection of forest plots. If substantial heterogeneity was detected (*I^2^* > 50%) a random effects model was employed; otherwise the data were analysed using a fixed effects model.

Relative risk (RR) estimates for dichotomous outcomes and weighted mean differences (MD) for continuous outcomes were calculated with corresponding 95% confidence intervals.

Statistical significance was determined based on an alpha level of 0.05.

## 3. Results

### 3.1 Study selection

A total of 298 articles were returned by the primary literature search. One further article was identified via screening of the reference lists of previous reviews. Removal of duplicates resulted in 243 unique articles. Of these, 220 articles were excluded following screening of titles and abstracts. The full texts of the remaining 23 articles were reviewed, resulting in five eligible studies being identified (Figure 1).

**Figure 1.**
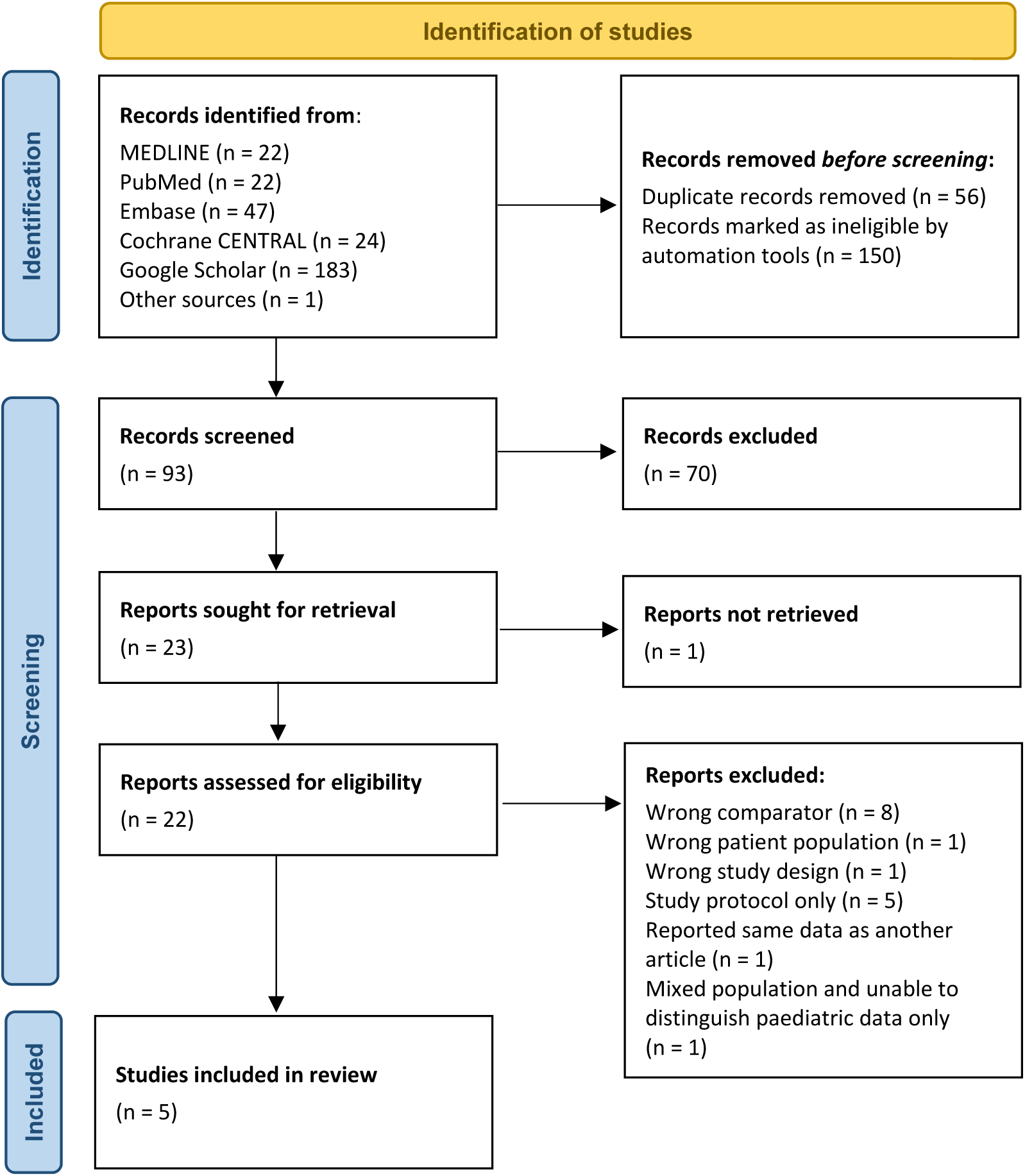
Prisma diagram

### 3.2 Study characteristics

Of the five studies [29–33], three [29, 30, 32] compared intramuscular midazolam with buccal midazolam (Table 1). One of these studies also included an intranasal midazolam treatment arm [30]. The remaining studies [31, 33] compared intramuscular midazolam with rectal diazepam. All studies included children treated in the hospital emergency department by hospital medics. The study in which intramuscular midazolam was compared with both buccal and intranasal midazolam also recruited a home group in which the emergency ASM was administered by the children’s parents. There were no studies in which data for emergency ASM administration by paramedics were reported.

**Table 1.** Characteristics of included studies.

| Study | Country | Design | Setting | Inclusion criteria | Exclusion criteria | Outcome measures | IM midazolam group | Comparator group(s) |
| --- | --- | --- | --- | --- | --- | --- | --- | --- |
| Alansari (2020)<br>NCT02897856<br>[29] | Qatar | Double-blind,<br>double-dummy,<br>randomised trial | General hospital<br>paediatric<br>emergency centre | Age 6 mths to 14 years;<br>Active convulsive seizure<br>(abnormal motor activity<br>with associated loss of<br>consciousness on<br>presentation) lasting > 5<br>minutes;<br>No intravenous ASM<br>access.<br>Status epilepticus defined<br>as continuous or<br>interrupted abnormal<br>motor activity with loss of<br>consciousness and no<br>cessation of seizure<br>activity for ≥ 30 minutes | Cardiac arrest or<br>hemodynamic instability<br>at presentation;<br>Seizure associated with<br>head trauma or drowning;<br>confirmed diagnosis of<br>bacterial meningitis or<br>viral/bacterial<br>encephalitis; Significant<br>hypoglycaemia or<br>electrolyte abnormality;<br>History of congenital heart<br>disease, inborn error of<br>metabolism or allergy to<br>benzodiazepines | Seizure cessation<br>(physician-confirmed<br>cessation of abnormal<br>motor activity with at least<br>partial recovery of<br>consciousness) within 5<br>minutes of study drug<br>administration (primary);<br>Need for additional ASM<br>treatment;<br>Duration of seizure activity;<br>seizure cessation within 10<br>minutes of study drug<br>administration;<br>Continuation of active<br>seizure for ≥ 30 minutes<br>after arrival at emergency<br>dept;<br>Recurrence of seizure<br>activity within 1 hour;<br>Length of hospital stay;<br>PICU admission; Adverse<br>effects | N (randomised) = 75<br>N (analysed) = 67<br>Dose 0.05 mL/kg (max 1.6<br>mL)/0.3 mg/kg (max 8<br>mg) | Buccal midazolam<br>N (randomised) = 75<br>N (analysed) = 70<br>Dose 0.06 mL/kg (max 2<br>mL)/0.25 mg/kg (max 10<br>mg) |
| Mohammed<br>(2024)<br>NCT05670509<br>[30] | Egypt | Unblinded,<br>randomised<br>controlled trial | Hospital<br>emergency dept.;<br>Patients' home | Age 1 mth to 17 years;<br>Known seizure disorder;<br>Active convulsive seizure<br>and ≥ 3 convulsions<br>within preceding hour OR<br>active convulsive seizure<br>and ≥ 2 convulsions in<br>succession without<br>recovery of consciousness<br>OR active single<br>convulsive seizure lasting<br>≥ 5 minutes | Non-convulsive status<br>epilepticus;<br>Benzodiazepine treatment<br>within 1 hour of<br>presentation;<br>Known history of<br>hypersensitivity, non-<br>responsiveness or other<br>contraindication to<br>benzodiazepines;<br>Use of concomitant drugs<br>contraindicated in<br>combination with<br>benzodiazepines;<br>Significant hypotension,<br>cardiac dysrhythmia or<br>current hypoglycaemia on<br>presentation to hospital; | Cessation<br>(caregiver/physician-confirmed absence of<br>abnormal motor activity<br>with partial or full recovery<br>of consciousness) of visible<br>seizure activity within 10<br>minutes with sustained<br>absence of visible seizure<br>activity for 30 minutes<br>following single ASM dose<br>(primary);<br>Time to seizure cessation;<br>Time taken for ASM<br>preparation and<br>administration | N (randomised; ED) = 34<br>N (analysed; ED) = 34<br>N (randomised; Home) =<br>69<br>N (analysed; Home) = NR<sup>1</sup><br>Dose 0.2 mg/kg (max 10<br>mg) | Buccal midazolam<br>N (randomised; ED) = 37<br>N (analysed; ED) = 37<br>N (randomised; Home) =<br>67<br>N (analysed; Home) = NR<sup>1</sup><br>Dose 0.2 mg/kg (max 10<br>mg)<br>Intranasal midazolam<br>N (randomised; ED) = 34<br>N (analysed; ED) = 34<br>N (randomised; Home) =<br>60<br>N (analysed; Home) = NR<sup>1</sup><br>Dose 0.2 mg/kg (max (10<br>mg) |
|  |  |  |  |  | Caregiver objection to use of randomised intervention |  |  |  |
| Momen (2015) [31] | Iran | Unblinded, randomised controlled trial | Tertiary children's hospital emergency dept. | Age $\geq 1$ mth; Convulsive status epilepticus (continuous generalised convulsive seizure lasting $\geq 5$ minutes) at time of presentation at emergency department | Established intravenous access on arrival at hospital; Administration of rectal or nasal benzodiazepines by parents or paramedics; Failure of parents to give verbal consent to participate; Serial seizures without recovery of consciousness between seizures; History of serious adverse reaction to study medications | Cessation of convulsive seizure activity within 10 minutes of ASM administration without seizure recurrence within 60 minutes of initial seizure cessation (primary); Time to cessation of seizure from time of ASM administration; Adverse effects | N (randomised) = 50<br>N (analysed) = 50<br>Dose 0.3 mg/kg | Rectal diazepam<br>N (randomised) = 50<br>N (analysed) = 50<br>Dose 0.5 mg/kg |
| Muchohi (2008) [32] | Kenya | Open-label, non-randomised, uncontrolled trial | Hospital paediatric high dependency unit | Age 6 mths to 13 years; Features of severe malaria; Convulsion lasting $\geq 5$ minutes or $\geq 3$ convulsions lasting $< 5$ minutes within 1 hour; Parental written informed consent | Treatment with diazepam before admission to ED; Compromised cardio-respiratory function; Detectable midazolam concentrations in pre-dose blood sample; Inability to obtain parental informed consent | Cessation of visible signs of seizure activity within 10 minutes of ASM administration without respiratory depression or seizure recurrence within 6 hours (primary); Time to seizure cessation from time of ASM administration; Seizure recurrence; Respiratory depression; Death | N = 12<br>Dose 0.3 mg/kg | Buccal midazolam<br>N = 8<br>Dose 0.3 mg/kg |
| Munir (2019) [33] | Pakistan | Open-label, randomised trial | Hospital paediatric department | Age 1 mth to 18 years; Admission to hospital with any acute seizure type; Parental informed consent | History of allergy to benzodiazepines; Respiratory depression; Acute diarrhoea; Status epilepticus | Time to seizure cessation from time of ASM administration; Cessation of seizure within 5 minutes of ASM administration | N (randomised) = 33<br>N (analysed) = 33<br>Dose 0.2 mg/kg | Rectal diazepam<br>N (randomised) = 33<br>N (analysed) = 33<br>Dose 0.5 mg/kg |

**Abbreviations:** ASM = Antiseizure medication; ED = Emergency department; NR = Not reported; PICU = Paediatric intensive care unit
**Notes:** (1). Home data reported as number of seizure episodes. Some participants provided data for $> 1$ seizure.

Four studies were randomised [29–31, 33], one of which used a double-blind, double-dummy design to compare intramuscular midazolam with buccal midazolam [29]. The remaining study was open-label and non-randomised [32]. In total, 640 participants were enrolled of whom 444 received treatment in the emergency department and 196 received treatment at home. Data were reported for 613 participants, 428 treated in the emergency department and 185 treated at home. Of the 428 treated in the emergency department, 196 received intramuscular midazolam, 34 intranasal midazolam, 115 buccal midazolam and 83 rectal diazepam. In the single study reporting data for children treated at home [30], some participants in the at-home group provided data for more than one seizure episode, resulting in data being reported for a total of 196 seizures in 185 individuals of which 69 seizures were treated with intramuscular midazolam, 60 with intranasal midazolam and 67 with buccal midazolam. Per-protocol midazolam doses, irrespective of route of administration, were between 0.2 mg/kg and 0.3 mg/kg. Intramuscular midazolam was administered at a dose of 0.3 mg/kg in three of the five studies [29, 31, 32] and 0.2 mg/kg in the remaining studies [30, 33]. The maximum per-protocol dose of intramuscular midazolam was 8 mg in one study [29], 10 mg in one study [30] and was not stated in the remaining studies [31–33]. Rectal diazepam was administered at a dose of 0.5 mg/kg [31, 33] (Table 1).

All studies described treatment for acute seizures but seizure eligibility criteria varied between studies: four studies included individuals with single convulsive seizures lasting ≥ 5 minutes [29–32]; two studies additionally included individuals with ≥ 3 convulsive seizures within the preceding hour [30, 32]; and one study additionally included individuals with ≥ 2 convulsive seizures in succession without recovery of consciousness [30]. The remaining study included individuals with any acute seizure but excluded individuals presenting with status epilepticus [33]. The studies were conducted in Qatar, Egypt, Iran, Kenya and Pakistan.

### 3.3 Efficacy outcomes

The main efficacy outcomes for each of the included studies are summarised in Table 2.

**Table 2.**
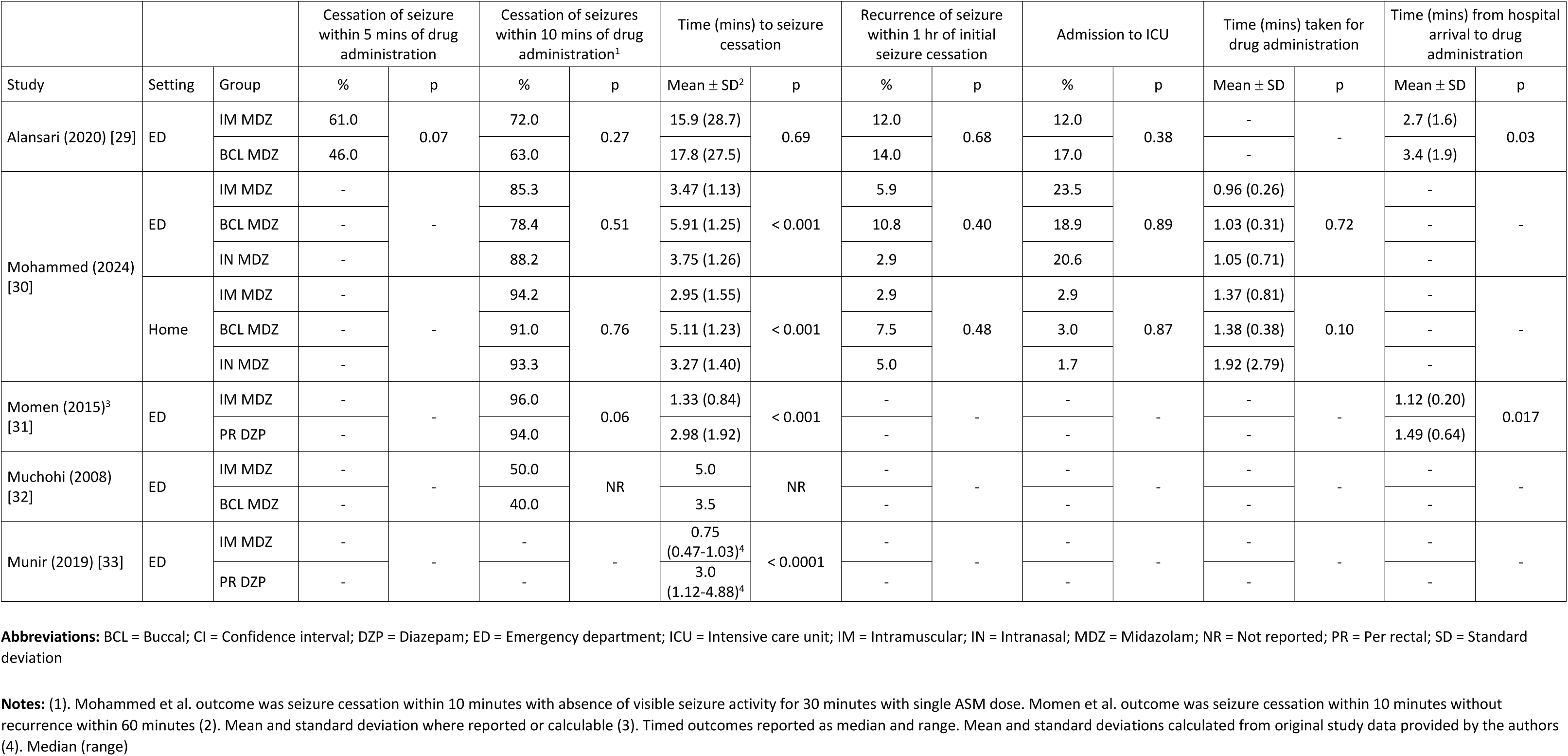
Main efficacy findings for included studies.

|  |  |  | Cessation of seizure within 5 mins of drug administration |  | Cessation of seizures within 10 mins of drug administration<sup>1</sup> |  | Time (mins) to seizure cessation |  | Recurrence of seizure within 1 hr of initial seizure cessation |  | Admission to ICU |  | Time (mins) taken for drug administration |  | Time (mins) from hospital arrival to drug administration |  |
| --- | --- | --- | --- | --- | --- | --- | --- | --- | --- | --- | --- | --- | --- | --- | --- | --- |
| Study | Setting | Group | % | p | % | p | Mean ± SD<sup>2</sup> | p | % | p | % | p | Mean ± SD | p | Mean ± SD | p |
| Alansari (2020) [29] | ED | IM MDZ | 61.0 | 0.07 | 72.0 | 0.27 | 15.9 (28.7) | 0.69 | 12.0 | 0.68 | 12.0 | 0.38 | - | - | 2.7 (1.6) | 0.03 |
|  |  | BCL MDZ | 46.0 |  | 63.0 |  | 17.8 (27.5) |  | 14.0 |  | 17.0 |  | - |  | 3.4 (1.9) |  |
| Mohammed (2024) [30] | ED | IM MDZ | - | - | 85.3 | 0.51 | 3.47 (1.13) | < 0.001 | 5.9 | 0.40 | 23.5 | 0.89 | 0.96 (0.26) | 0.72 | - | - |
|  |  | BCL MDZ | - |  | 78.4 |  | 5.91 (1.25) |  | 10.8 |  | 18.9 |  | 1.03 (0.31) |  | - |  |
|  |  | IN MDZ | - |  | 88.2 |  | 3.75 (1.26) |  | 2.9 |  | 20.6 |  | 1.05 (0.71) |  | - |  |
|  | Home | IM MDZ | - | - | 94.2 | 0.76 | 2.95 (1.55) | < 0.001 | 2.9 | 0.48 | 2.9 | 0.87 | 1.37 (0.81) | 0.10 | - | - |
|  |  | BCL MDZ | - |  | 91.0 |  | 5.11 (1.23) |  | 7.5 |  | 3.0 |  | 1.38 (0.38) |  | - |  |
|  |  | IN MDZ | - |  | 93.3 |  | 3.27 (1.40) |  | 5.0 |  | 1.7 |  | 1.92 (2.79) |  | - |  |
| Momen (2015)<sup>3</sup> [31] | ED | IM MDZ | - | - | 96.0 | 0.06 | 1.33 (0.84) | < 0.001 | - | - | - | - | - | - | 1.12 (0.20) | 0.017 |
|  |  | PR DZP | - |  | 94.0 |  | 2.98 (1.92) |  | - |  | - |  | - |  | 1.49 (0.64) |  |
| Muchohi (2008) [32] | ED | IM MDZ | - | - | 50.0 | NR | 5.0 | NR | - | - | - | - | - | - | - | - |
|  |  | BCL MDZ | - |  | 40.0 |  | 3.5 |  | - |  | - |  | - |  | - |  |
| Munir (2019) [33] | ED | IM MDZ | - | - | - | - | 0.75 (0.47-1.03)<sup>4</sup> | < 0.0001 | - | - | - | - | - | - | - | - |
|  |  | PR DZP | - |  | - |  | 3.0 (1.12-4.88)<sup>4</sup> |  | - |  | - |  | - |  | - |  |

**Abbreviations:** BCL = Buccal; CI = Confidence interval; DZP = Diazepam; ED = Emergency department; ICU = Intensive care unit; IM = Intramuscular; IN = Intranasal; MDZ = Midazolam; NR = Not reported; PR = Per rectal; SD = Standard deviation
**Notes:** (1). Mohammed et al. outcome was seizure cessation within 10 minutes with absence of visible seizure activity for 30 minutes with single ASM dose. Momen et al. outcome was seizure cessation within 10 minutes without recurrence within 60 minutes (2). Mean and standard deviation where reported or calculable (3). Timed outcomes reported as median and range. Mean and standard deviations calculated from original study data provided by the authors (4). Median (range)

#### 3.3.1 Seizure cessation within 5-10 minutes of drug administration

In individual studies, seizure cessation within 10 minutes of drug administration was similar for intramuscular, buccal and intranasal midazolam [29, 30], and for intramuscular midazolam compared with rectal diazepam [31]. Point estimates tended to favour intramuscular midazolam. In the study by Alansari et al. [29], seizure cessation within 10 minutes was reported in 72% in the intramuscular midazolam group compared with 63% in the buccal midazolam group. It should be noted that seizures that had not responded to initial benzodiazepine treatment within 5 minutes in this study were treated with additional ASMs; however, seizure cessation at 5 minutes following initial benzodiazepine administration also numerically favoured intramuscular midazolam (61% vs 46%; p = 0.07). In the study by Mohammed et al. [30], caregivers reported seizure cessation within 10 minutes in a higher percentage of those treated at home relative to those treated in hospital. 94.2% of children treated at home with intramuscular midazolam had seizure cessation compared with 93.3% treated with intranasal midazolam and 91.0% treated with buccal midazolam. In the emergency department groups in the same study, the corresponding proportions were 85.3%, 88.2% and 78.4%. Muchohi et al. [32] reported seizure cessation within 10 minutes in 50% treated with intramuscular midazolam and 40% treated with buccal midazolam. Finally, in the study by Momen et al. [31], 96% of individuals treated with intramuscular midazolam had cessation of seizures within 10 minutes compared with 94% treated with rectal diazepam.

Meta-analysis was possible only for the comparison of intramuscular midazolam with buccal midazolam. There was no difference between treatment groups in the analysis including data for both the emergency department and home settings (RR = 1.07; 95% CI = 0.97-1.17; Z = 1.40; p = 0.16) (Figure 2A) or analysis of data for the emergency department only (RR = 1.17; 95% CI = 0.97-1.40; Z = 1.69; p = 0.09) (Figure 3A). However, point estimates favoured intramuscular midazolam. The results of the meta-analysis are heavily influenced by a single study and that study is rated as having high overall risk of bias [30]. Since the protocol for the Alansari et al. study stipulated additional ASM treatment for seizures that had not been controlled within 5 minutes of initial benzodiazepine administration, the 5-minute data for this study were used in the meta-analysis for this outcome. For the other studies included in the analysis, data were for seizure cessation within 10 minutes.

**Figure 2.**
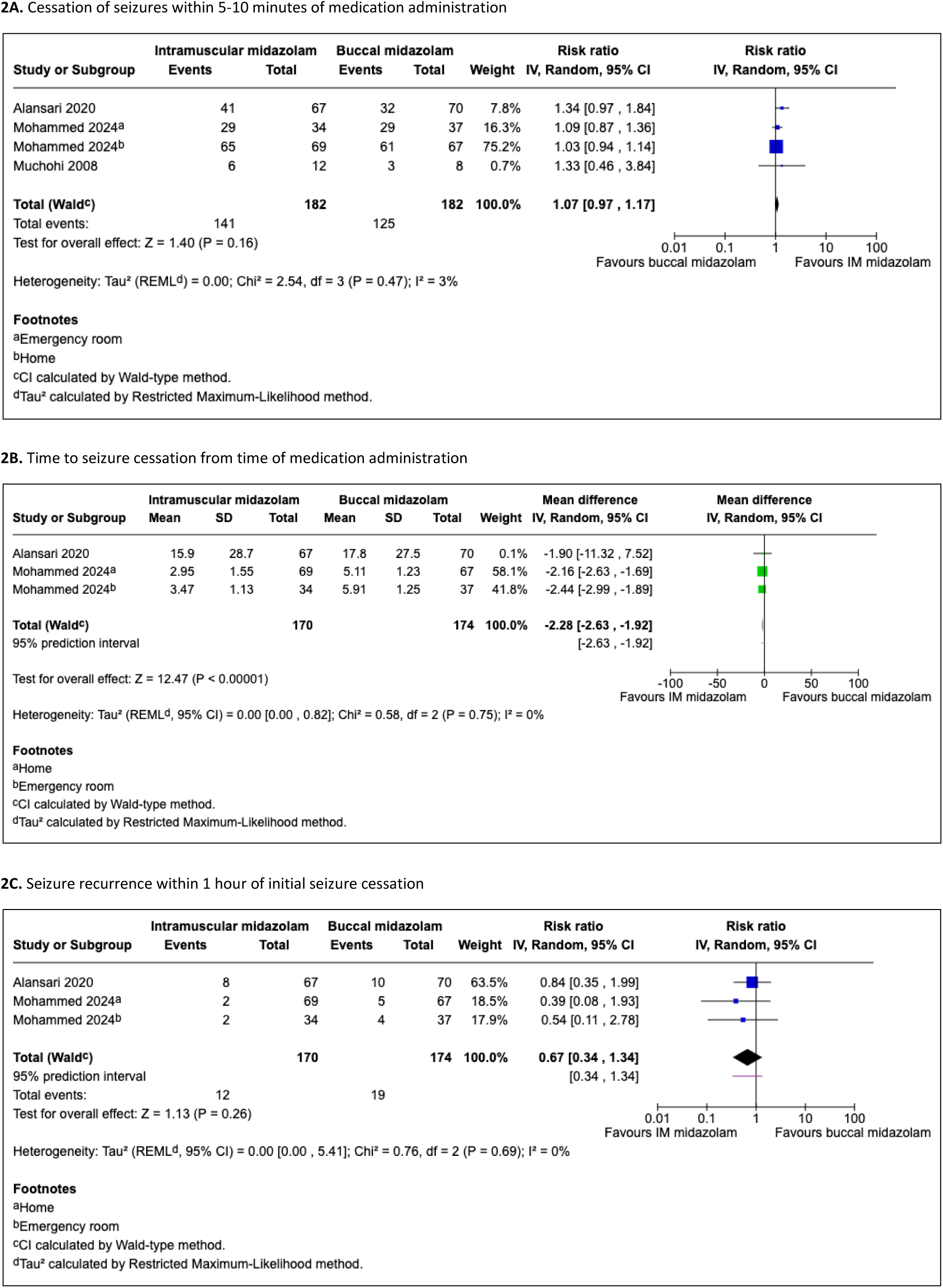
Efficacy outcomes for intramuscular midazolam vs buccal midazolam (data for emergency room and home groups)

**Figure 3.**
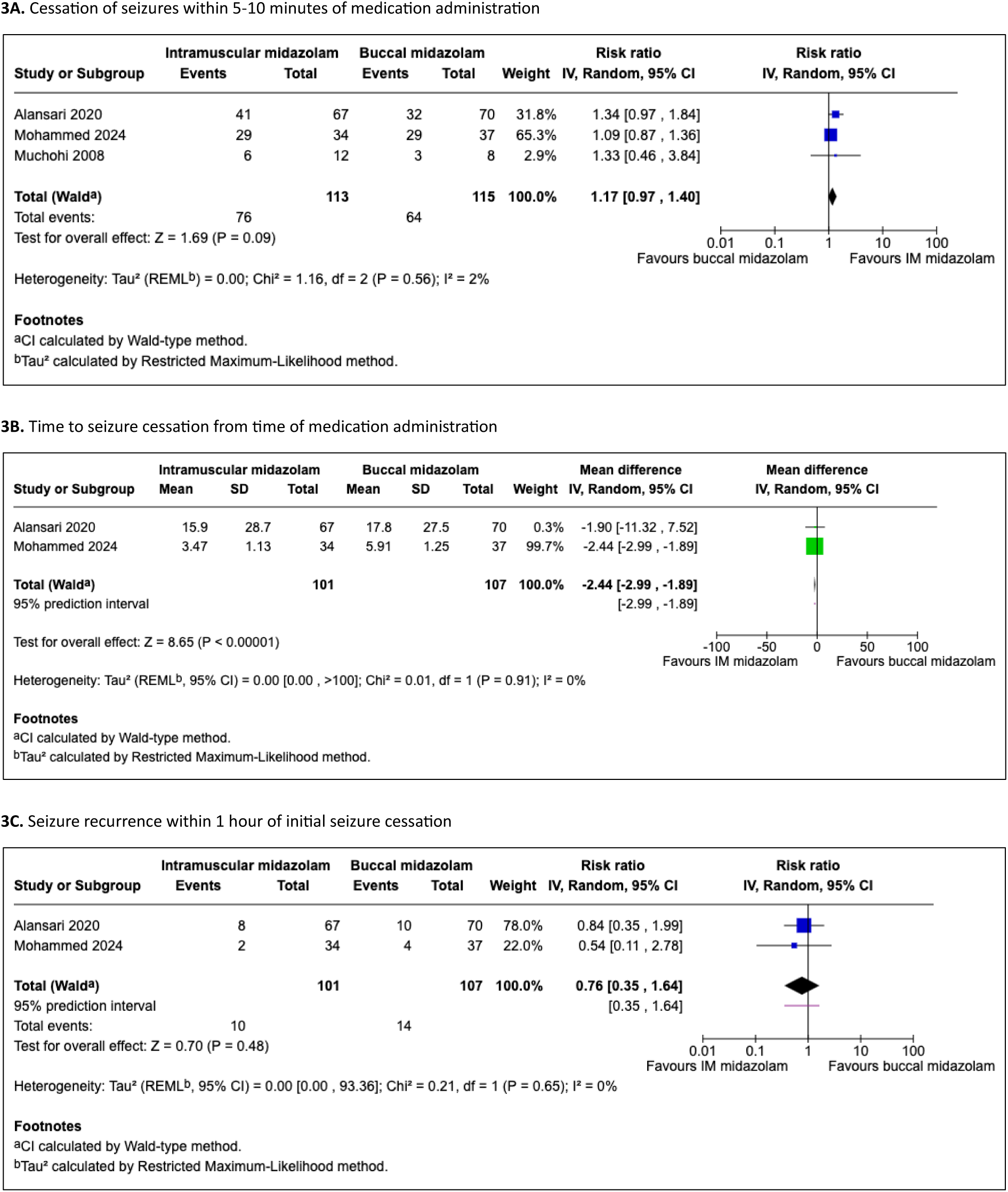
Efficacy outcomes for intramuscular midazolam vs buccal midazolam (data for emergency department only)

#### 3.3.2 Time to seizure cessation after drug administration

In individual studies, time to seizure cessation after drug administration was shorter with intramuscular midazolam than with intranasal midazolam or buccal midazolam in both home and emergency department treatment groups in one study (both p < 0.0001) [30], but there was no difference between intramuscular and buccal midazolam in another study (p = 0.69), although point estimates favoured intramuscular midazolam (15.9 ± 28.7 vs 17.8 ± 27.5 mins) [29]. The median time to seizure cessation in the study in children with seizures associated with malaria was longer for intramuscular midazolam (5 mins) than for buccal midazolam (3.5 mins) [32]. In the studies comparing intramuscular midazolam to rectal diazepam, time to seizure cessation was shorter in the intramuscular midazolam group (p < 0.001) [31, 33].

In the meta-analysis, intramuscular midazolam was superior to buccal midazolam in analysis, including data for both emergency department and home groups (MD = −2.28; 95% CI: −2.63 to −1.92; Z = 12.47; p < 0.0001) (Figure 2B) and data for the emergency department only (MD = −2.44; 95% CI: −2.99 to −1.89; Z = 8.65; p < 0.0001) (Figure 3B).

#### 3.3.3 Recurrence of seizures within 1 hour of initial seizure cessation

In individual studies, there was no difference in seizure recurrence within one hour between intramuscular midazolam, intranasal midazolam or buccal midazolam [29, 30], or between intramuscular midazolam and rectal diazepam [31]. Point estimates typically, but not always, favoured intramuscular midazolam: 12% vs 14% for buccal midazolam in the study by Alansari et al. [29]; 2.9%, 7.5% and 5.0% for intramuscular, buccal and intranasal routes, respectively, in the home groups, and 5.9%, 10.8% and 2.9% for intramuscular, buccal and intranasal routes, respectively, in the emergency room groups in the study by Mohammed et al. [30]. In the study comparing intramuscular midazolam with rectal diazepam, there was no recurrence of seizures within one hour in any of the individuals who had cessation of the index seizure within 10 minutes of drug administration in either treatment group [31].

In the meta-analysis, there was no difference between intramuscular midazolam and buccal midazolam either in the analysis including data for both emergency department and home settings (RR = 0.67; 95% CI = 0.34-1.34; Z = 1.13; p = 0.26) (Figure 2C) or the analysis of data for the emergency department only (RR = 0.76; 95% CI = 0.35-1.64; Z = 0.70; p = 0.48) (Figure 3C).

### 3.4 Incidence of respiratory depression and hypotension

Very few cases of respiratory depression or hypotension were reported in any of the studies. In one study, a single child developed respiratory depression and hypotension three minutes after administration of intramuscular midazolam and was successfully treated with oral airway, bag valve mask ventilation and 20 mL/kg bolus of normal saline [29]. In another study, respiratory depression was reported in a child mistakenly administered double the intended dose of intramuscular midazolam (0.6 mg/kg) who recovered following bag valve mask ventilation [31]. Cases of respiratory depression were also reported in children with seizures associated with severe malaria treated with either intramuscular midazolam (n = 1) or buccal midazolam (n = 2) all of which were successfully treated with the benzodiazepine antagonist flumazenil [32]. The study excluded children who had compromised cardio-respiratory function at the time of admission to the paediatric high dependency unit (but not those in respiratory distress) but did not exclude children who were admitted having already entered coma. However, no information was provided on respiration or level of consciousness before midazolam administration in the children who were recruited and provided outcome data.

This study was the only study in which deaths were reported (one in the intramuscular midazolam group and three in the buccal midazolam group), all of which occurred at least 24 hours after midazolam administration and were attributed to complications associated with malaria [32].

### 3.5 Risk of bias

Only two of the five studies [29, 33] were considered to have a low overall risk of bias (Figure 4). The other three studies [30–32] were considered to have a high risk of bias overall and specifically in relation to randomisation (one study was not randomised), concealment of treatment allocation and blinding of outcome assessors.

**Figure 4.**
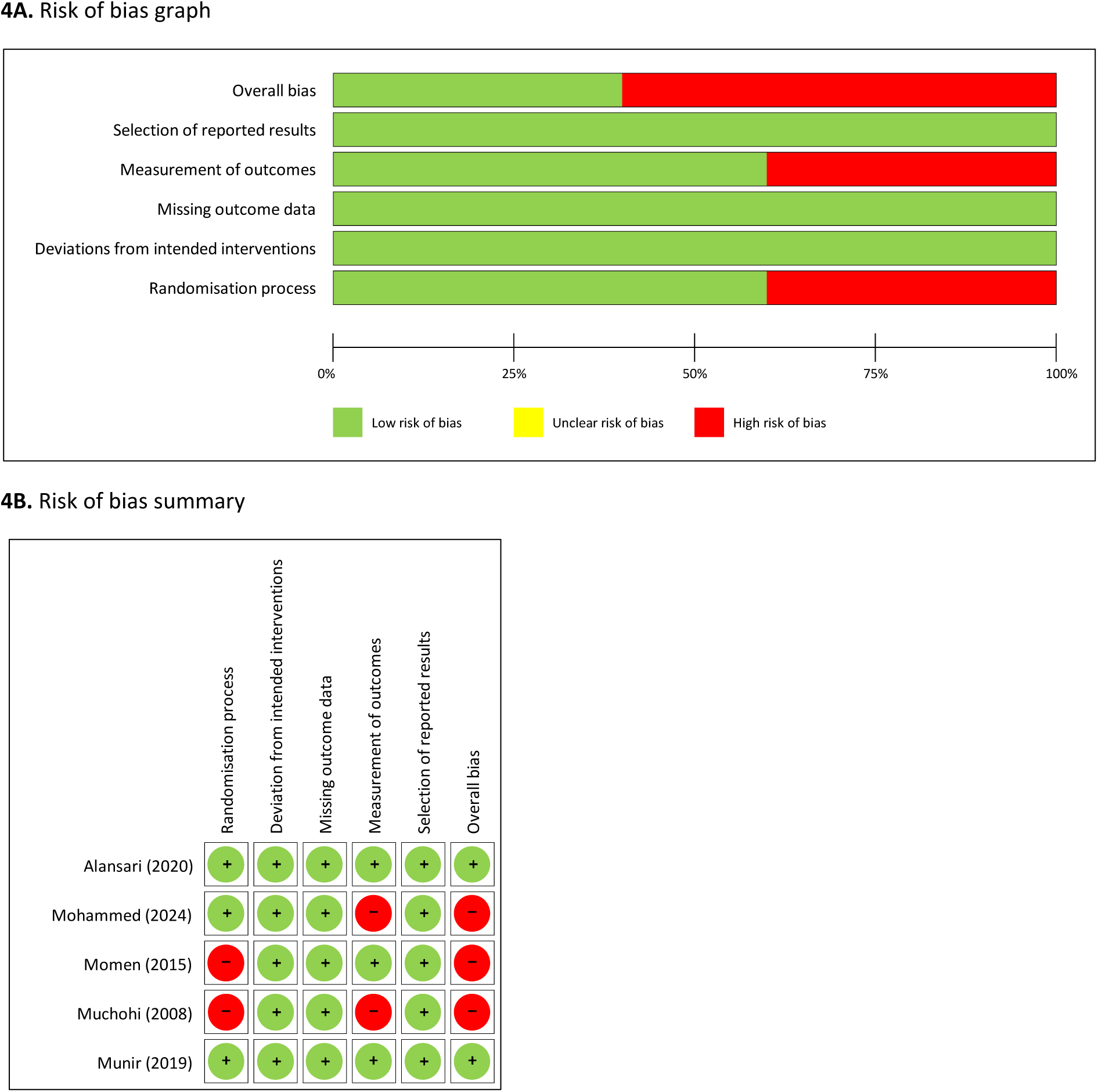
Risk of bias

### 3.6 Grade of evidence

For the primary outcome, seizure cessation within 5-10 minutes of medication administration, the quality of evidence was “low” supporting the finding of no difference in effectiveness between intramuscular midazolam and buccal midazolam, and “very low” for no difference in effectiveness between intramuscular midazolam and intranasal midazolam (supplementary material). Seizure cessation within 5-10 minutes was not reported in either of the studies comparing intramuscular midazolam with rectal diazepam.

## 4. Discussion

Data from the five studies identified in this review suggest that intramuscular midazolam may have similar safety and efficacy to intranasal midazolam, buccal midazolam or rectal diazepam for the treatment of acute paediatric seizures in direct comparisons. However, the quality of evidence was rated as low or very low, and paediatric data from randomised studies comparing intramuscular midazolam and other non-intravenous benzodiazepines are scarce. Further, the results of meta-analyses comparing intramuscular versus buccal midazolam are heavily influenced by a single study and that study is rated as having high overall risk of bias.

Previous systematic reviews have tended to include data for intravenous lorazepam and intravenous diazepam implying that the conclusions may be less relevant to out-of-hospital and pre-hospital settings. Since meta-analyses have also often relied on grouping of data for midazolam irrespective of route of administration, the relative safety and efficacy of intramuscular midazolam compared with midazolam by other routes and other non-intravenous benzodiazepines remains to be ascertained. However, the broader analyses have consistently found that midazolam by any non-intravenous route has at least comparable efficacy to intravenous or non-intravenous diazepam and to intravenous lorazepam in terms of successful seizure cessation and total time to seizure cessation (including time take for drug administration) [20, 24, 34–37]. In addition, network meta-analyses suggest intramuscular midazolam compares favourably to other non-intravenous benzodiazepines for seizure outcomes.

For example, a recent network meta-analysis [34] of data for children treated with benzodiazepines either in the hospital emergency department or by paramedics found intramuscular midazolam was associated with the highest mean absolute risk and best rank probability (indicating the highest probability of being the most effective treatment) for number of successful seizure cessations.

In an earlier network meta-analysis of predominantly paediatric data from open-label randomised trials conducted in hospital emergency departments [38] intramuscular midazolam was superior to other non-intravenous medications, including rectal diazepam, for time to seizure cessation after administration, time to seizure cessation after arrival at hospital and time to initiate treatment.

Given the imperative to abort the seizure as rapidly as possible, time from drug administration to seizure cessation is the primary metric of effectiveness where all other considerations are equal; however, it is also evident that time taken for administration and ease of administration vary markedly between administration routes and are particularly important considerations where drugs are administered in out-of-hospital settings and by non-medical personnel.

A related concern is confidence in administering the medication; rectal administration tends to be the least preferred option with patients, caregivers and emergency services personnel, largely because of perceived stigma and concerns over social acceptability [19, 39, 40].

In the study by Mohammed et al. [30] comparing intramuscular, intranasal and buccal midazolam, caregivers expressed greatest satisfaction with the intranasal route whereas hospital clinicians favoured the intramuscular route. Among caregivers, 30% considered intramuscular administration to be “very easy” or “easy” compared with 87% for intranasal and 90% for buccal administration. The corresponding figures for the clinician group were 97%, 91% and 97%, respectively, for intramuscular, intranasal and buccal routes. In both groups, the mean time to seizure cessation was shortest with intramuscular midazolam, suggesting the perceived relative difficulty with intramuscular administration in the caregiver group was not an impediment to successful treatment.

In 2022, a single-dose, prefilled autoinjector form of midazolam was licensed in the United States for the treatment of status epilepticus in adults [39, 41]. The use of intramuscular midazolam for paediatric seizures in prehospital settings is now a recommendation in treatment protocols in the United States [42, 43] but this is despite there being lack of clinical trial evidence for intramuscular midazolam for out of hospital or prehospital treatment of paediatric CSE. An injectable form of midazolam which can be administered either intravenously or intramuscularly is licensed for sedation and anaesthesia, but not for CSE [44].

## 5. Limitations

The findings of the current review should be interpreted cautiously given the various limitations of the data and the low quality of evidence. The small number of studies and paucity of data for most comparisons limited the feasibility of a comprehensive data synthesis. Only a single study provided data for the comparison of intramuscular midazolam with intranasal midazolam. No studies were identified comparing intramuscular midazolam with intranasal diazepam. Meta-analysis was only possible for the comparison of intramuscular midazolam with buccal midazolam. Inconsistencies in methodology, sample characteristics, reporting and outcomes mean the results may not be generalisable to the broader population of children presenting with CSE. Weight-based doses of intramuscular midazolam were higher in three out of the five studies (0.3 mg/kg) than the typically recommended dose for children (0.2 mg/kg). While it is unclear whether this might have affected seizure outcomes, there were very few reported cases of respiratory depression or hypotension suggesting that the higher doses were relatively well tolerated. Only one study included a treatment group in which initial rescue medication for seizures was administered outside of the hospital emergency department. We were unable to find any studies comparing intramuscular midazolam to other non-intravenous benzodiazepines in the prehospital ambulance setting. Since CSE frequently occurs in previously neurologically typical children implying that prehospital treatment in many cases will be administered by paramedics, this represents an important shortcoming in the available data. Few data were available on the duration of seizures at the time of admission to hospital in those who received initial intervention in the emergency department meaning the time taken to initiate treatment is unclear in most cases. Since none of the studies included long-term follow-ups, it was not possible to investigate the relationship between time to intervention and the risk of long-term adverse outcomes.

## 6. Conclusions

The evidence from randomised clinical trials is insufficient to draw firm conclusions with regard to the effectiveness of intramuscular midazolam relative to that of other non-intravenous benzodiazepines for the treatment of acute paediatric seizures. Limited data from individual randomised comparisons suggest intramuscular midazolam may have comparable safety and efficacy to midazolam administered by intranasal or buccal routes, and to rectally administered diazepam, but the quality of evidence is low or very low. There are very few data for out-of-hospital administration of intramuscular midazolam compared to other non-intravenous benzodiazepines for paediatric seizures and no data from randomised studies for out-of-hospital or prehospital treatment by emergency service personnel. Further studies, in particular for out-of-hospital and prehospital settings, are warranted. Consistent inclusion criteria, definition of eligible seizure episodes, outcome measures and method of ascertainment should be used to facilitate future data syntheses.

## Declarations

### III. Availability of data and material

Not applicable.

### IV. Ethics approval

Not applicable.

### V. Consent to participate

Not applicable.

### VI. Consent for publication

Not applicable.

### VII. Code availability

Not applicable.

### VIII. Author contributions

Literature search, systematic review and data analysis conducted by RC and MV. All authors contributed to the preparation of the manuscript and approved the final version for submission.

## Data Availability

There are no original data associated with this review

