## Supplementary file for "A systematic review of efficacy and safety of intramuscular midazolam versus midazolam by other routes or other non-intravenous benzodiazepines in children with convulsive status epilepticus"

1. Search strategy (MEDLINE)

2. Summary of findings table (Intramuscular midazolam vs buccal midazolam)

3. Summary of findings table (Intramuscular midazolam vs intranasal midazolam)

**1. Search strategy (MEDLINE)**

1. exp Epilepsy, Tonic-Clonic/
2. (tonic adj2 clonic).tw
3. exp Seizures/
4. epilep$.tw
5. seizure$.tw
6. convuls$.tw
7. exp Status Epilepticus/
8. (status adj2 epilepticus).tw
9. 1 or 2 or 3 or 4 or 5 or 6 or 7 or 8
10. randomized controlled trial.pt
11. controlled clinical trial.pt
12. pragmatic clinical trial.pt
13. clinical trial.pt
14. (clin$ adj2 (study or trial)).tw
15. ((singl$ OR doubl$ or trebl$ or trip$) adj (blind$ or mask$)).tw
16. (control$ adj2 (study or trial)).tw
17. randomi$.tw
18. (randomi$ adj (allocate$ or assign$)).tw
19. exp Random Allocation/
20. exp Double-Blind Method/
21. exp Single-Blind Method/
22. exp Clinical Trial/
23. 10 or 11 or 12 or 13 or 14 or 15 or 16 or 17 or 18 or 19 or 20 or 21 or 22
24. 9 and 23
25. exp Infant/
26. exp Child/

27. exp Adolescent/

28. (pediatr$ or paediat$ or child$ or adolesc$).tw

29. 25 or 26 or 27 or 28

30. 24 and 29

31. ((intramuscul$ or IM) adj2 (midazolam)).tw

32. 30 and 31

33. (1980:3000/12/12).pdat

34. 32 and 33

35. (animals not human).sh

36. 34 not 35

**2. Summary of findings:**

**Intramuscular midazolam compared to buccal midazolam for acute seizures or convulsive status epilepticus in children and young people**

**Patient or population:** acute seizures or convulsive status epilepticus in children and young people

**Setting:** Out of hospital, pre-hospital or hospital emergency department

**Intervention:** intramuscular midazolam

**Comparison:** buccal midazolam

| Outcome<br>№ of participants<br>(studies) | Relative effect<br>(95% CI) | Anticipated absolute effects (95% CI) | | | Certainty | What happens |
|---|---|---|---|---|---|---|
| | | **buccal midazolam** | **intramuscular midazolam** | **Difference** | | |
| Cessation of seizure within 5-10 minutes of medication administration (Seizure cessation within 5-10 mins)<br>№ of participants: 364<br>(3 RCTs) | **RR 1.07**<br>(0.97 to 1.17) | 68.7% | **73.5%**<br>(80.4 to 66.6) | **4.8% more**<br>(2.1 fewer to 11.7 more) | ⊕⊕◯◯<br>Low[a,b] | The evidence suggests that intramuscular midazolam results in little to no difference in cessation of seizures within 5-10 minutes of medication administration compared with buccal midazolam. |

***The risk in the intervention group** (and its 95% confidence interval) is based on the assumed risk in the comparison group and the **relative effect** of the intervention (and its 95% CI).

**CI:** confidence interval; **RR:** risk ratio

**GRADE Working Group grades of evidence**
**High certainty:** we are very confident that the true effect lies close to that of the estimate of the effect.
**Moderate certainty:** we are moderately confident in the effect estimate: the true effect is likely to be close to the estimate of the effect, but there is a possibility that it is substantially different.
**Low certainty:** our confidence in the effect estimate is limited: the true effect may be substantially different from the estimate of the effect.
**Very low certainty:** we have very little confidence in the effect estimate: the true effect is likely to be substantially different from the estimate of effect.

**Explanations**

a. Two of three studies included in analysis accounting for > 90% of weighting considered to have high risk of bias

b. Limted data for outcome from small pooled sample size (< 380) and small total number of events (< 300)

**3. Summary of findings:**

**Intramuscular midazoam compared to intranasal midazolam for acute seizures or convulsive status epilepticus in young people**

**Patient or population:** acute seizures or convulsive status epilepticus in young people

**Setting:** out of hospital, pre-hospital or hospital emergency department

**Intervention:** intramuscular midazoam

**Comparison:** intranasal midazolam

| Outcome<br>№ of participants<br>(studies) | Relative effect<br>(95% CI) | Anticipated absolute effects (95% CI) | | | Certainty | What happens |
|---|---|---|---|---|---|---|
| | | **intranasal midazolam** | **intramuscular midazoam** | **Difference** | | |
| Seizure cessation within 5-10 minutes of medication administration (Seizure cessation within 5-10 mins)<br>№ of participants: 197<br>(1 RCT) | **RR 1.03**<br>(0.41 to 2.55) | 91.5% | **94.2%**<br>(100 to 37.5) | **2.7% more**<br>(54 fewer to 141.8 more) | ⊕◯◯◯<br>Very low[a,b] | Very limted evidence suggests that intramuscular midazolam results in little to no difference in cessation of seizures within 5-10 minutes of medication administration compared with intranasal midazolam. |

***The risk in the intervention group** (and its 95% confidence interval) is based on the assumed risk in the comparison group and the **relative effect** of the intervention (and its 95% CI).

**CI:** confidence interval; **RR:** risk ratio

**GRADE Working Group grades of evidence**
**High certainty:** we are very confident that the true effect lies close to that of the estimate of the effect.
**Moderate certainty:** we are moderately confident in the effect estimate: the true effect is likely to be close to the estimate of the effect, but there is a possibility that it is substantially different.
**Low certainty:** our confidence in the effect estimate is limited: the true effect may be substantially different from the estimate of the effect.
**Very low certainty:** we have very little confidence in the effect estimate: the true effect is likely to be substantially different from the estimate of effect.

**Explanations**

a. Single study judged to be at high risk of bias

b. Single study with small sample size (< 200) and small number of events (< 200)
